# Group testing: revisiting the ideas

**DOI:** 10.1101/2020.06.29.20142323

**Authors:** Viktor Skorniakov, Remigijus Leipus, Gediminas Juzeliūnas, Kęstutis Staliūnas

## Abstract

The task of identification of randomly scattered ‘bad’ items in a fixed set of objects is a frequent one, and there are many ways to deal with it. ‘Group testing’ (GT) refers to the testing strategy aiming to effectively replace the inspection of single objects by the inspection of groups spanning more than one object. First announced by Dorfman in 1943, the methodology has underwent vigorous development, and though many related research still takes place, the ground ideas remain the same. In the present paper, we revisit two classical GT algorithms: the Dorfman’s algorithm and the halving algorithm. Our fresh treatment of the latter and expository comparison of the two is devoted to dissemination of GT ideas which are so important in the current COVID-19 induced pandemic situation.

## 1 Introduction

The task of identification of bad items in a given set of objects arises quite often. For example, consider identification of: a) the infected patients in a fixed cohort; or b) the defective items in the production batch. Usually, this identification task is a composite problem and spans many subtasks. One of such subtasks can be described as ‘an efficient utilization of resources devoted to testing of investigated objects’. It turns out that, among plenitude of context dependent methods designed for the solution of this subtask, the appropriately chosen testing plan plays an exceptional role since it alone can reduce the testing costs substantially. This is the contextual target of the present paper. To be more precise, we focus on testing strategies widely known under the name of Group (or Pooled Sample) Testing (in what follows, we make use of an abbreviation GT). The core idea underlying GT strategy is an observation that, in many cases, the testing of single items can be replaced by the testing of a group spanning more than one item. Though it is difficult to trace back the exact date and inventor of this cornerstone idea (for a good historical account see [13], Ch. 1), without doubt much of the credit goes to the pioneering work of Dorfman [12]. In that paper, the Blood Testing problem was described and the following scheme was suggested. Given *N* individual blood samples, pool them and test for the presence of an infection in the pooled sample; in case of the negative test – finish; in case of the positive test – retest each single patient. The rationale behind is clear: if the prevalence of the infection is low, one usually ends up with a single test applied to the pool instead of *N* tests applied individually.

Since appearance of Dorfman’s work [12] in 1943, GT ideas were evolving in many directions and found important applications in molecular biology, quality control, computer science and other fields. Digging into the literature, one can observe that it is indeed very widespread across different disciplines. Because of this reason, some developments were overlapping and rediscovered by researchers working in the different fields. Our personal familiarity with the field also underwent this route: attracted by potential applications in the context of COVID-19 epidemics, we have rediscovered some well known facts. Nonetheless, the attained experience and understanding of the importance of the tool inspired us to write a promotional paper on the topic. This is the main intent of the paper: we believe that, in the current pandemic situation, the spread of GT ideas and attraction of other researchers to the field is an important and meaningful task. We do not propose novel GT schemes or methodological improvements. Our presentation is primarily devoted to those unfamiliar with the subject aiming to provide a quick lightweight introduction ‘by example’ without delving into details yet giving a flavor of the topic as a whole. Choosing a mathematical journal we, first of all, were interested in the dissemination within the *mathematically oriented* community. Secondly, while getting familiar with the topic, we have encountered a lot of papers where the subject was treated without sufficient mathematical rigorousness. We therefore felt that our rigorous treatment of the GT Scheme **H** (see Section 2), unseen (or at least unobserved) by us, was a missing item in the existing literature. Finally, after submission of the initial version of the paper, we have discovered that our Proposition 2.2 adds some new information to what is known about classical Dorfman’s scheme (see the comments in Section 2).

The remaining part of the paper is organized as follows. In Section 2, we provide some preliminaries, then describe and contrast two classical GT schemes. In Section 3, we give an accompanying discussion highlighting some relevant issues and skim through the related literature. Appendices A and B contain some mathematical derivations and tables. Because of COVID and the exemplary nature, we attach the whole presentation to the biomedical context.

## 2 Two classical GT schemes

Consider the following setup. Assume that the prevalence of some disease (the fraction of the infected individuals) is equal to *p* ∈ (0, 1) in an infinite (or large enough) population. A cohort spanning *N* independent individuals has to be tested and infected patients have to be identified. To achieve the goal, samples are collected from each individual. The applied test performs equally well for individual and for pooled samples: a situation might occur, e.g., when the test indicates the presence of the infection in the blood sample and there is no difference whether the latter is obtained from a single individual or from a pooled cohort of samples. For the situation described, physicians can choose different testing strategies. Let us assume that the following are three possible choice†.

**Scheme A:** Test each patient’s sample.

**Scheme D:** Conduct testing of the pooled sample. Test each member of the cohort separately only in case of detected infection in the pooled sample.

**Scheme H:**

*Step 1*. Test pooled sample of the whole cohort. Proceed to *Step 2*.

*Step 2*. If the test is positive, proceed to

*Step 3*. otherwise, finish testing cohort. *Step 3*. Divide the cohort into two parts consisting of the first and second halves respectively. Apply the whole algorithm to the two obtained parts recursively.

Although it is not obvious at the first glance, schemes **D** and **H** can be much more efficient as compared to **A** provided prevalence *p* is low enough. To give a rigorous justification (along with the concept of *efficiency*), let us formally define the underlying model.

Consider the sample of *N* individuals. Put *X*_*i*_ = 1, provided the test of *i*-th individual is positive, and *X*_*i*_ = 0, otherwise. Let *S* = *S*_*N*_ = *X*_1_ + … + *X*_*N*_ be the total number of infected individuals in the sample and let *T* = *T*_*N*_ be the total number of tests applied to the cohort.

We start with the scheme **D**. The test is applied once if the result is negative, and it is further applied to each of *N* individuals otherwise, i.e.

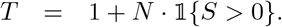

The above implies that *X*_1_, *…, X*_*N*_ are independent identically distributed (i.i.d.) random variables each having Bernoulli distribution Be(*p*). Therefore, *S* has the Binomial distribution Bin(*N, p*). Consequently, an average number of tests per cohort is

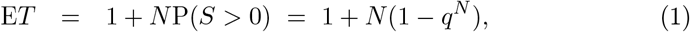

where *q* := 1 − *p*. An average number of tests per individual, say *t* = *t*(*N*), is

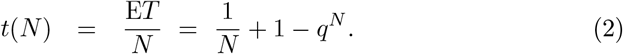

Consider a function *t* : (0, ∞) → (0, ∞) given in (2). By equating its derivative to 0, we see that the stationary points solve equation

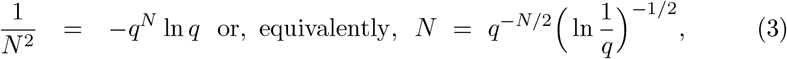

which is a fixed point equation for *g*(*N*) = *q*^−*N/*2^ ln(1*/q*) ^−1/2^, *N* ∈ (0, ∞), and, hence, can be easily solved iteratively. It is further not difficult to prove that, for *p* in the region enclosing (0, 0.2), there exists a unique solution *N*_*p*_ *>* 0 of (3) which is a minimizer of *t*(*N*) (see Proposition 2.2 below). Then, turning back to economic/biomedical interpretation, we conclude that, having a cohort of ⌊*N*_*p*_ ⌋ (here and in the sequel, ⌊ *y* ⌋ ∈ *ℝ* stands for an integer part of *y* individuals, Scheme **D** results in a lowest average number of tests per person which is possible when applying scheme of this type for a population having prevalence *p*. Scheme **A**, in contrast, always has a constant number of tests 1 per person. Therefore, an average (absolute) gain attained applying Scheme **D** instead of Scheme **A** is given by the difference

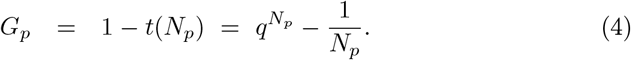

Right panel in Figure 1 shows the graph of *p ↦* 100 *G*_*p*_, *p* ∈ (0, 0.2), which is an average gain measured by the number of tests saved per 100 individuals. The corresponding values are provided in Table 1 (see Appendix B). An accompanying graph of *p* ↦ *N*_*p*_ (see the left panel of the Figure 1) demonstrates dependence of an optimal sample size on *p*. To obtain a fast numerical evidence, assume that *N* is bounded away from zero and *pN* → 0. Then from (3) it follows that the optimal sample size satisfies

**Table 1:** Performance of Scheme **D**

| $N_p$ | Range of $p$ | Range of $100G_p$ | $N_p$ | Range of $p$ | Range of $100G_p$ |
| --- | --- | --- | --- | --- | --- |
| 2 | 0.1865 – 0.2000 | 14.0000 – 16.1782 | 22 | 0.0020 – 0.0021 | 90.9350 – 91.1457 |
| 3 | 0.0855 – 0.1864 | 20.5225 – 43.1472 | 23 | 0.0019 – 0.0019 | 91.3723 – 91.3723 |
| 4 | 0.0506 – 0.0854 | 44.9721 – 56.2450 | 24 | 0.0017 – 0.0018 | 91.6016 – 91.8321 |
| 5 | 0.0336 – 0.0505 | 57.1747 – 64.2917 | 25 | 0.0016 – 0.0016 | 92.0759 – 92.0759 |
| 6 | 0.0239 – 0.0335 | 64.8434 – 69.8233 | 26 | 0.0015 – 0.0015 | 92.3261 – 92.3261 |
| 7 | 0.0179 – 0.0238 | 70.1977 – 73.8374 | 27 | 0.0014 – 0.0014 | 92.5843 – 92.5843 |
| 8 | 0.0140 – 0.0178 | 74.1163 – 76.8337 | 28 | 0.0013 – 0.0013 | 92.8517 – 92.8517 |
| 9 | 0.0112 – 0.0139 | 77.0523 – 79.2489 | 29 | 0.0012 – 0.0012 | 93.1296 – 93.1296 |
| 10 | 0.0091 – 0.0111 | 79.4383 – 81.2637 | 30 | 0.0011 – 0.0011 | 93.4188 – 93.4188 |
| 11 | 0.0076 – 0.0090 | 81.4428 – 82.8596 | 32 | 0.0010 – 0.0010 | 93.7241 – 93.7241 |
| 12 | 0.0065 – 0.0075 | 83.0288 – 84.1396 | 33 | 0.0009 – 0.0009 | 94.0421 – 94.0421 |
| 13 | 0.0055 – 0.0064 | 84.2998 – 85.3889 | 35 | 0.0008 – 0.0008 | 94.3806 – 94.3806 |
| 14 | 0.0048 – 0.0054 | 85.5569 – 86.3428 | 38 | 0.0007 – 0.0007 | 94.7426 – 94.7426 |
| 15 | 0.0042 – 0.0047 | 86.5106 – 87.2152 | 41 | 0.0006 – 0.0006 | 95.1303 – 95.1303 |
| 16 | 0.0037 – 0.0041 | 87.3879 – 87.9915 | 45 | 0.0005 – 0.0005 | 95.5524 – 95.5524 |
| 17 | 0.0033 – 0.0036 | 88.1708 – 88.6533 | 50 | 0.0004 – 0.0004 | 96.0195 – 96.0195 |
| 18 | 0.0030 – 0.0032 | 88.8385 – 89.1800 | 58 | 0.0003 – 0.0003 | 96.5507 – 96.5507 |
| 19 | 0.0027 – 0.0029 | 89.3683 – 89.7296 | 71 | 0.0002 – 0.0002 | 97.1814 – 97.1814 |
| 20 | 0.0024 – 0.0026 | 89.9265 – 90.3079 | 100 | 0.0001 – 0.0001 | 98.0049 – 98.0049 |
| 21 | 0.0022 – 0.0023 | 90.5176 – 90.7183 |  |  |  |

**Figure 1.**
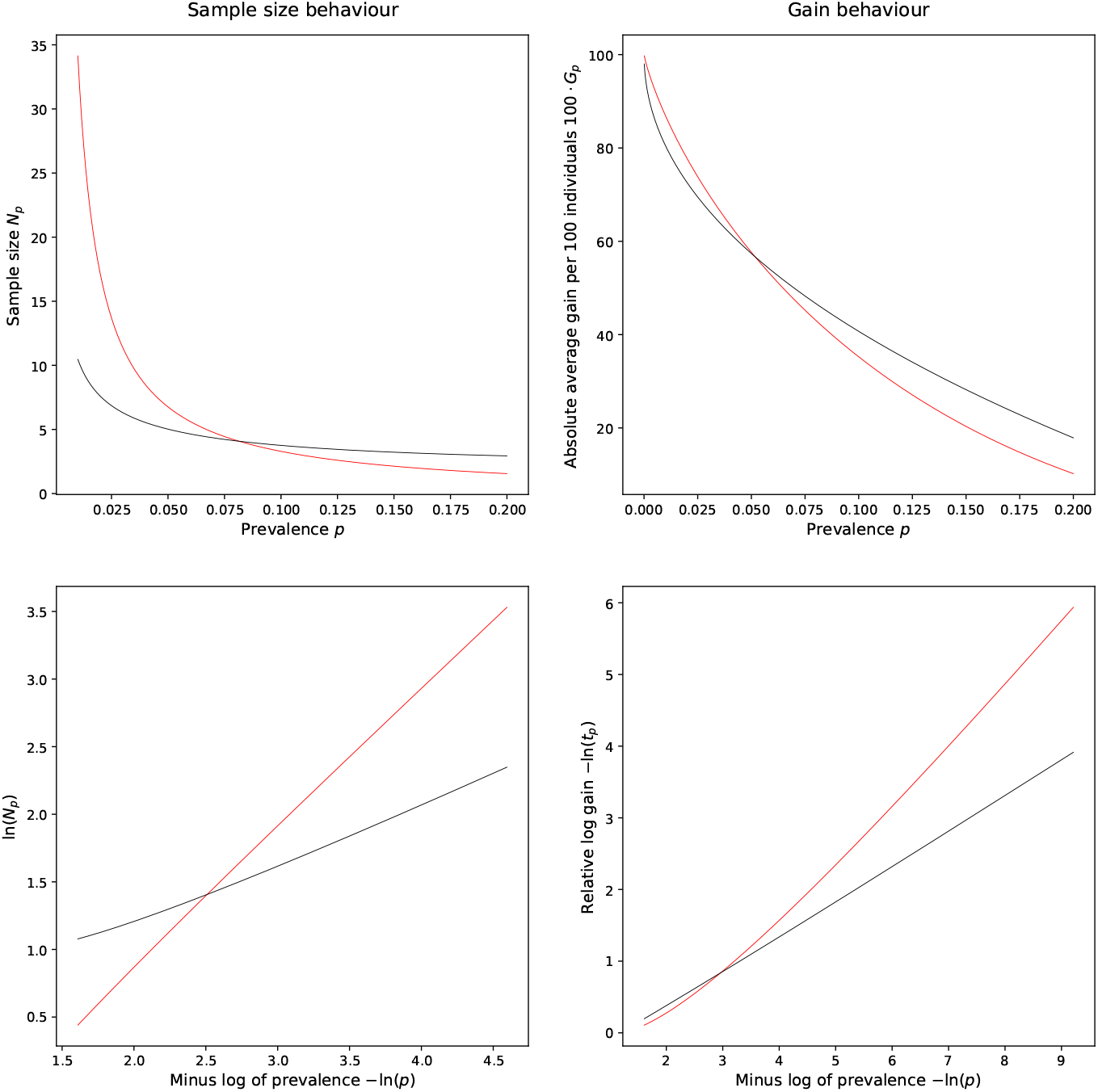
Scheme **H** (red) vs. Scheme **D** (black) on the linear and the log-log scale.

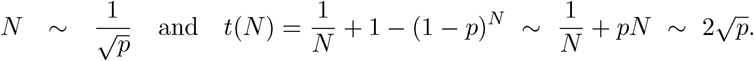

Hence, assuming that *p* is small enough for *pN* ≈ 0 to hold, the above implies that

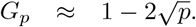

For example, if *p* = 0.01, then we have *G*_*p*_ ≈ 0.8, i.e. an approximate average gain is 80% or so.

Now let us switch to the Scheme **H**. Its main features are summarized in the following proposition (for the proof see Appendix A).

### Proposition 2.1.

*Assume the Scheme* ***H***. *Then*

i. *an average number of tests per person is given by*

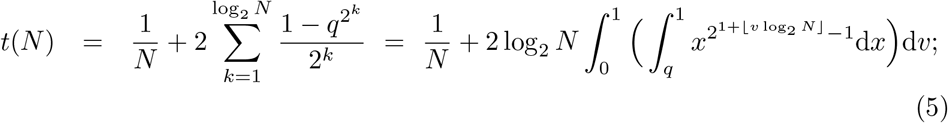
ii. *an average number of tests per person in the case of an infinitely large cohort is*

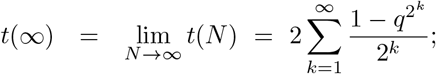
iii. *for a fixed p* ∈ (0, 1), *function t* : ℕ *1* ↦ (0, ∞) *admits at most two minimizers N*_*p*_: *the value N* = *N*_*p*_ *corresponding to optimal sample size is either* 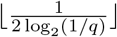 *or* 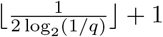.

Inspection of the results in the statement of the proposition leads to a quick comparison of Scheme **H** with **A** and **D**. Indeed, consider first the limit in (ii). Obviously,

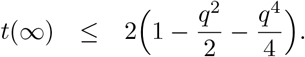

Hence, for *q* ≈ 1 (or alternatively *p* ≈ 0), *t*(∞) < 1. The latter means that, when the prevalence is low, this scheme always outperforms common sequential Scheme **A**. Again, to gain a quick quantitative insight, assume that *p* is small enough for *pN* ≈ 0 to hold. Then turning to (iii) and taking a ‘continuous’ (undiscretized) version of *N*_*p*_ equal to 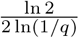 yields relationships (see Remark A.1, eq. (9))

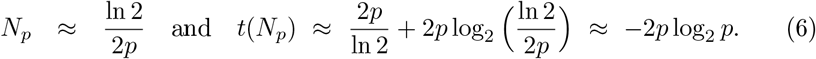

Therefore, an approximation to an average gain *G*_*p*_ = 1 − *t*(*N*_*p*_) is 1 + 2*p* log_2_ *p*. Taking, e.g., *p* = 0.01 results in *G*_0.01_ ≈ 0.867. Considering analogous example given for Scheme **D**, we see that the gain has an increase close to 7%. In fact, this is not surprising (for a visual comparison of Schemes **D** and **H** on the linear and the log-log scales, see Figure 1, and, for the numerical one, Table 1 and Table 2 in Appendix B) since, for Scheme **D**, we had 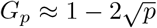 and 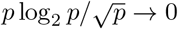 as *p* → 0. Equality (6), however, exhibits some magic flavor. To see this, note that, for *p* ≈ 0, entropy *I*_*p*_ of *X ∼* Be(*p*) is asymptotically equivalent to *p* log_2_ *p* since

**Table 2:** Performance of Scheme **H**

| $N_p$ | Range of $p$ | Range of $100G_p$ | $N_p$ | Range of $p$ | Range of $100G_p$ |
| --- | --- | --- | --- | --- | --- |
| 1 | 0.1592 – 0.2000 | 10.2068 – 18.1573 | 59 | 0.0058 – 0.0058 | 91.4813 – 91.4813 |
| 2 | 0.1092 – 0.1591 | 18.1801 – 32.0493 | 60 | 0.0057 – 0.0057 | 91.5996 – 91.5996 |
| 3 | 0.0830 – 0.1091 | 32.0828 – 41.7999 | 61 | 0.0056 – 0.0056 | 91.7184 – 91.7184 |
| 4 | 0.0670 – 0.0829 | 41.8413 – 48.8858 | 62 | 0.0055 – 0.0055 | 91.8377 – 91.8377 |
| 5 | 0.0562 – 0.0669 | 48.9333 – 54.2777 | 64 | 0.0054 – 0.0054 | 91.9575 – 91.9575 |
| 6 | 0.0484 – 0.0561 | 54.3303 – 58.5384 | 65 | 0.0053 – 0.0053 | 92.0779 – 92.0779 |
| 7 | 0.0424 – 0.0483 | 58.5953 – 62.0600 | 66 | 0.0052 – 0.0052 | 92.1987 – 92.1987 |
| 8 | 0.0378 – 0.0423 | 62.1207 – 64.9241 | 67 | 0.0051 – 0.0051 | 92.3202 – 92.3202 |
| 9 | 0.0341 – 0.0377 | 64.9881 – 67.3443 | 69 | 0.0050 – 0.0050 | 92.4422 – 92.4422 |
| 10 | 0.0311 – 0.0340 | 67.4112 – 69.3911 | 70 | 0.0049 – 0.0049 | 92.5648 – 92.5648 |
| 11 | 0.0285 – 0.0310 | 69.4607 – 71.2322 | 72 | 0.0048 – 0.0048 | 92.6880 – 92.6880 |
| 12 | 0.0264 – 0.0284 | 71.3044 – 72.7691 | 73 | 0.0047 – 0.0047 | 92.8118 – 92.8118 |
| 13 | 0.0245 – 0.0263 | 72.8434 – 74.2009 | 75 | 0.0046 – 0.0046 | 92.9362 – 92.9362 |
| 14 | 0.0229 – 0.0244 | 74.2775 – 75.4396 | 76 | 0.0045 – 0.0045 | 93.0612 – 93.0612 |
| 15 | 0.0215 – 0.0228 | 75.5181 – 76.5498 | 78 | 0.0044 – 0.0044 | 93.1869 – 93.1869 |
| 16 | 0.0202 – 0.0214 | 76.6301 – 77.6042 | 80 | 0.0043 – 0.0043 | 93.3132 – 93.3132 |
| 17 | 0.0191 – 0.0201 | 77.6863 – 78.5153 | 82 | 0.0042 – 0.0042 | 93.4402 – 93.4402 |
| 18 | 0.0181 – 0.0190 | 78.5990 – 79.3593 | 84 | 0.0041 – 0.0041 | 93.5678 – 93.5678 |
| 19 | 0.0172 – 0.0180 | 79.4445 – 80.1325 | 86 | 0.0040 – 0.0040 | 93.6962 – 93.6962 |
| 20 | 0.0164 – 0.0171 | 80.2193 – 80.8312 | 88 | 0.0039 – 0.0039 | 93.8253 – 93.8253 |
| 21 | 0.0157 – 0.0163 | 80.9193 – 81.4518 | 91 | 0.0038 – 0.0038 | 93.9552 – 93.9552 |
| 22 | 0.0150 – 0.0156 | 81.5412 – 82.0814 | 93 | 0.0037 – 0.0037 | 94.0858 – 94.0858 |
| 23 | 0.0144 – 0.0149 | 82.1721 – 82.6285 | 96 | 0.0036 – 0.0036 | 94.2172 – 94.2172 |
| 24 | 0.0138 – 0.0143 | 82.7204 – 83.1829 | 98 | 0.0035 – 0.0035 | 94.3493 – 94.3493 |
| 25 | 0.0133 – 0.0137 | 83.2760 – 83.6506 | 101 | 0.0034 – 0.0034 | 94.4824 – 94.4824 |
| 26 | 0.0128 – 0.0132 | 83.7448 – 84.1237 | 104 | 0.0033 – 0.0033 | 94.6162 – 94.6162 |
| 27 | 0.0124 – 0.0127 | 84.2190 – 84.5063 | 108 | 0.0032 – 0.0032 | 94.7509 – 94.7509 |
| 28 | 0.0119 – 0.0123 | 84.6025 – 84.9897 | 111 | 0.0031 – 0.0031 | 94.8866 – 94.8866 |
| 29 | 0.0115 – 0.0118 | 85.0871 – 85.3808 | 115 | 0.0030 – 0.0030 | 95.0231 – 95.0231 |
| 30 | 0.0112 – 0.0114 | 85.4792 – 85.6767 | 119 | 0.0029 – 0.0029 | 95.1607 – 95.1607 |
| 31 | 0.0108 – 0.0111 | 85.7759 – 86.0750 | 123 | 0.0028 – 0.0028 | 95.2992 – 95.2992 |
| 32 | 0.0105 – 0.0107 | 86.1752 – 86.3764 | 128 | 0.0027 – 0.0027 | 95.4388 – 95.4388 |
| 33 | 0.0102 – 0.0104 | 86.4775 – 86.6804 | 133 | 0.0026 – 0.0026 | 95.5794 – 95.5794 |
| 34 | 0.0099 – 0.0101 | 86.7822 – 86.9868 | 138 | 0.0025 – 0.0025 | 95.7211 – 95.7211 |
| 35 | 0.0096 – 0.0098 | 87.0896 – 87.2959 | 144 | 0.0024 – 0.0024 | 95.8640 – 95.8640 |
| 36 | 0.0094 – 0.0095 | 87.3996 – 87.5035 | 150 | 0.0023 – 0.0023 | 96.0081 – 96.0081 |
| 37 | 0.0091 – 0.0093 | 87.6078 – 87.8172 | 157 | 0.0022 – 0.0022 | 96.1534 – 96.1534 |
| 38 | 0.0089 – 0.0090 | 87.9224 – 88.0279 | 164 | 0.0021 – 0.0021 | 96.3001 – 96.3001 |
| 39 | 0.0087 – 0.0088 | 88.1337 – 88.2398 | 173 | 0.0020 – 0.0020 | 96.4481 – 96.4481 |

Table 2 – *Continued from previous page*
| $N_p$ | Range of $p$ | Range of $100G_p$ | $N_p$ | Range of $p$ | Range of $100G_p$ |
| --- | --- | --- | --- | --- | --- |
| 40 | 0.0085 – 0.0086 | 88.3463 – 88.4532 | 182 | 0.0019 – 0.0019 | 96.5976 – 96.5976 |
| 41 | 0.0083 – 0.0084 | 88.5603 – 88.6678 | 192 | 0.0018 – 0.0018 | 96.7486 – 96.7486 |
| 42 | 0.0081 – 0.0082 | 88.7757 – 88.8839 | 203 | 0.0017 – 0.0017 | 96.9012 – 96.9012 |
| 43 | 0.0079 – 0.0080 | 88.9924 – 89.1014 | 216 | 0.0016 – 0.0016 | 97.0555 – 97.0555 |
| 44 | 0.0077 – 0.0078 | 89.2107 – 89.3203 | 230 | 0.0015 – 0.0015 | 97.2116 – 97.2116 |
| 45 | 0.0076 – 0.0076 | 89.4303 – 89.4303 | 247 | 0.0014 – 0.0014 | 97.3696 – 97.3696 |
| 46 | 0.0074 – 0.0075 | 89.5408 – 89.6515 | 266 | 0.0013 – 0.0013 | 97.5297 – 97.5297 |
| 47 | 0.0072 – 0.0073 | 89.7627 – 89.8743 | 288 | 0.0012 – 0.0012 | 97.6920 – 97.6920 |
| 48 | 0.0071 – 0.0071 | 89.9863 – 89.9863 | 314 | 0.0011 – 0.0011 | 97.8567 – 97.8567 |
| 49 | 0.0070 – 0.0070 | 90.0987 – 90.0987 | 346 | 0.0010 – 0.0010 | 98.0241 – 98.0241 |
| 50 | 0.0068 – 0.0069 | 90.2115 – 90.3247 | 384 | 0.0009 – 0.0009 | 98.1943 – 98.1943 |
| 51 | 0.0067 – 0.0067 | 90.4383 – 90.4383 | 433 | 0.0008 – 0.0008 | 98.3677 – 98.3677 |
| 52 | 0.0066 – 0.0066 | 90.5524 – 90.5524 | 494 | 0.0007 – 0.0007 | 98.5448 – 98.5448 |
| 53 | 0.0064 – 0.0065 | 90.6669 – 90.7819 | 577 | 0.0006 – 0.0006 | 98.7260 – 98.7260 |
| 54 | 0.0063 – 0.0063 | 90.8973 – 90.8973 | 692 | 0.0005 – 0.0005 | 98.9120 – 98.9120 |
| 55 | 0.0062 – 0.0062 | 91.0132 – 91.0132 | 866 | 0.0004 – 0.0004 | 99.1039 – 99.1039 |
| 56 | 0.0061 – 0.0061 | 91.1295 – 91.1295 | 1155 | 0.0003 – 0.0003 | 99.3030 – 99.3030 |
| 57 | 0.0060 – 0.0060 | 91.2463 – 91.2463 | 1732 | 0.0002 – 0.0002 | 99.5119 – 99.5119 |
| 58 | 0.0059 – 0.0059 | 91.3636 – 91.3636 | 3465 | 0.0001 – 0.0001 | 99.7360 – 99.7360 |

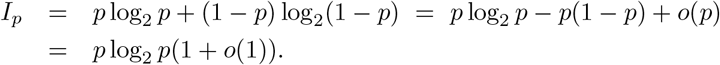

Consequently, (6) means that the optimal average number of tests per one individual scales like entropy of the prevalence of the infection. Keeping in a view the above relationship, it is not surprising that the significant number of works ([8], [19], [1]) have approached the testing problem from the Information Theory perspective. In the next section we provide additional comments regarding connections with the Information Theory. Here we end up with the previously mentioned Proposition 2.2, which is proved in Appendix A.

### Proposition 2.2.

*Let* 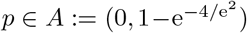 *be fixed. Consider function g*_*p*_(*N*) = *g*(*N*) = *q*^−*N/*2^(ln(1*/q*))^−1/2^, *N >* 0. *It admits a unique fixed point N*_*p*_ *which minimizes t*(*N*) *given by* (2).

The above proposition can be viewed as a counterpart of Proposition 2.1. Note, that it does not contain analytical expression of an optimal sample size. The latter was given by Samuels [35] and is either 1 + [*p*^−1/2^] or 2 + [*p*^−1/2^]. Of most importance is that Samuels [35] not only provided the analytical expression of the optimal sample size but has also shown that, for the case of Scheme **D**, the optimal sample size equals to 1 for *p >* 1 − (1/3)^1/3^ 0.31. This, in turn, is in agreement with the fundamental fact of the GT theory discovered by Ungar [42]: if 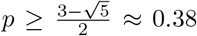, then there does not exist an algorithm that is better than individual one-by-one testing.

An interesting detail here is that our proof given in Appendix A differs from that of Samuels and leads to an exact analytical expression for the range of *p* (the set *A* above), where *g*_*p*_(*N*) has a unique minimizer.

## 3 Discussion

Since its appearance, the Dorfman’s Scheme **D** was rigorously investigated by many authors (we refer to [40], [39], [23], [35] to name a few). Talking about Scheme **H**, the situation is a bit different. To our best knowledge, the reference [46] is the only work close to ours both in nature of investigations and results. However, in that paper, the authors focus on the treatment of an asymptotic regime of Scheme **H** when *p* → 0. Majority of other references encountered by us provide instructions suitable for the practical application of Scheme **H** with a brief and nonrigorous theoretical background. For example, in the present context, it was currently afresh discussed by Gollier and Gossner [18], Mentus et al. [29] and Shani-Narkiss et al. [37]. For an older reference discussing the case of nonhomogeneous population (i.e., the one in which the probability of being infected *p* may vary across individuals) and containing quite a large body of applied literature on halving algorithm (i.e., Scheme **H**) we refer to [4].

One should have noticed that halving, constituting the core of the Scheme **H**, yields another link to the Information and Algorithm Theory in addition to the one already mentioned‡ in Section 2. Namely, in its essence, the Scheme **H** is nothing more than the Quick Sort (QS) algorithm designed to sort a set containing keys of two types (bad and good ones). It is well known that QS yields the best (up to the constant multiple) possible average performance among comparison-based algorithms: to sort an array having *N* non-constant (i.e., random) items, the smallest average number of comparisons is of the order *N* ln *N* [10], and all randomized ‘divide-and-conquer’ type algorithms (with QS being one among the rest) have expected time asymptotically equivalent to QS, which randomly splits sorted set into two equal subsets [11]. Our formula (5) is just a confirmation of the well known fact. To see this, note that, in the context of sorting task, (5) presents an average number of comparisons per item. Though the order is correct, we are inclined to think that the multiplier 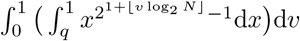 can be improved by making use of QS modification (or another comparison-based algorithm) designed to sort items with a small number of possible values (in our case, there are just two values: ‘sick’ and ‘healthy’). On the other hand, as already mentioned above, the order is optimal since though there are algorithms which can beat QS when sorting integers, e.g., [2], [41], they operate under different, i.e., noncomparison-based, mode. In our case, however, comparison is predefined by the setting of the problem at hand: we assume that biomedical tests can only be carried out by making use of comparison.

Though biomedical context is very frequent in applications, there are many others including engineering, environmental sciences, information theory, etc. (see [32], [21], [22], [30], [26], [15], [24], [28], [25], [6]). This ‘real life’ contextual diversity brings many constraints to take into account despite the fact that the standard binomial setting, considered in Section 2, quite often can be regarded as a good starting approximation. To get a full understanding of the matter touched, below is a shortlist of key issues with a brief description of each.

- Heterogeneity of population. The prevalence of disease may depend on other factors (e.g., age and gender).
- Imperfectness of the test. The test can have sensitivity and/or specificity below 1.
- Dilution effect. Pooling can reduce testing accuracy substantially. If this is the case, it is necessary to impose upper bound on the number of pooled samples.
- Implementation costs. In Section 2, we silently assumed that implementation of the considered schemes only involves retesting related costs. However, it may involve others as well.
- Dependence. It can happen that tested individuals are somehow related.

All these underpinnings have to be addressed carefully. Take, for example, the last one. From results presented in Section 2, one can infer that the application of the GT procedures is most effective when the prevalence is low (*p* ≈ 0). In such case, under classical assumption *pN* → *λ >* 0, the number of infected individuals *S*_*N*_ can be well approximated by the Poisson distribution Pois(*pN*), and the approximation remains quite accurate irrespectively of the nature of the dependence exhibited by summands (see [7], [43], [3] and references therein for results of this kind with possible extensions beyond the classical setting). It is therefore reasonable to assume that, after switching to Poisson approximation, at least some of the existing schemes can be carried over to the dependent case. Clearly, additional restrictions call for new theoretical investigations.

The set of directions of such investigations can be significantly appended by including other methods and GT related tasks. More concretely, the schemes considered in Section 2 broadly fall into the class of probabilistic GT schemes. Another widely adopted paradigm is called *combinatorial approach*. Within its framework, one does not assume any random mechanism and tries to make use of combinatorial methods in order to identify *d* bad items in the given group of *N* ≥ *d* objects (see monographs [13], [14]). Speaking about tasks, up to now, we have focused only on the identification of bad items (or infected patients) under assumption that the prevalence *p* is known. In addition to the literature devoted to this task, there is a huge body of literature dealing with an estimation (both point and interval) of *p* from pooled samples observations as well as testing issues (see, e.g., [34], [16], [20] and references therein).

We hope that our discussion complies well with our initial goal stated in the introduction. To emphasize the relevance of similar promotional discussions in the present context, we point out a huge burst of papers devoted to similar problems (see, e.g., [5], [17], [31], [33], [36], [38], [44], [45]). Besides that, we also note that some countries have already successfully applied pooling methodology for testing of the SARS-CoV-2 virus.§

## Data Availability

Data sharing is not applicable to this article as no new data were created or analysed in this study.

## A Appendix. Technical details

Proof of Proposition 2.1. (i) For 1 *≤ i ≤ j ≤ N* = 2^*n*^, let *M*_*ij*_ = {*X*_*i*_, *X*_*i*+1_, …, *X*_*j*_} and let 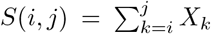 be the number of infected individuals in the cohort *M*_*ij*_. Let 𝕝_*i,j*_ denote an indicator function taking value 1 if there is at least one infected individual in the group *M*_*i,j*_, i.e. 𝕝 _i,j_ = 𝕝 *S*(*i, j*) *>* 0. Also, let *T* (*i, j*) be *the total number of tests applied to the cohort M*_*ij*_ *after the initial pooled test*. By the description of the testing Scheme **H**, applying recursive equations, we have

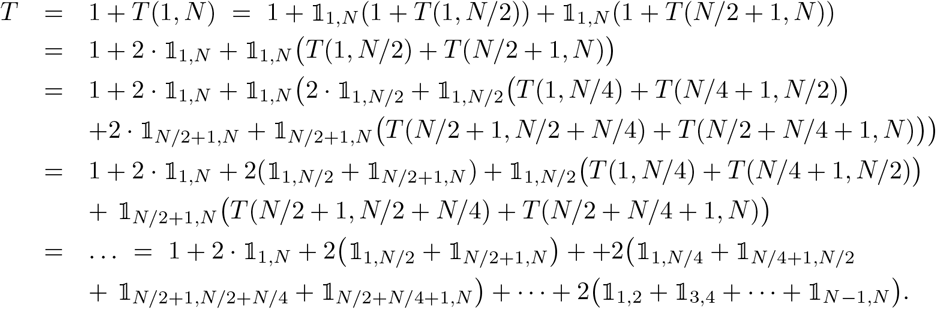

Taking expectations yields

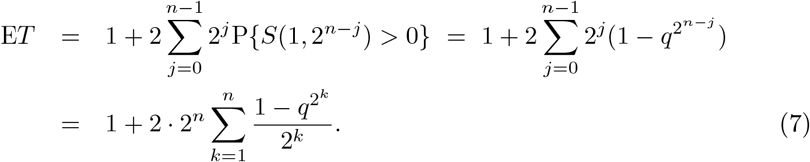

Hence, the first equality in

i. For the second one, take the last sum above and continue as follows:

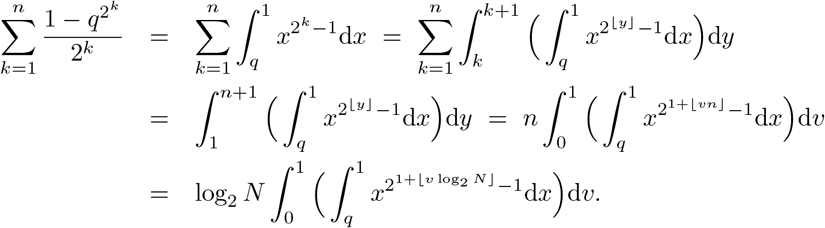
ii. By (7),

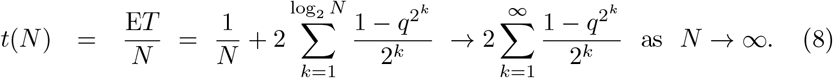
iii. Since *N* = *N* (*n*) = 2^*n*^, by the second equality in (8),

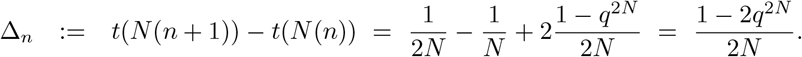

Clearly, *q*^2*N*^ → 0 as *N* → ∞. Therefore, there exist no more than two *N*_*p*_ ∈ *ℕ* such that Δ_*n*_ *≤* 0 for all *N ≤ N*_*p*_ and Δ_*n*_ *≥* 0 for all *N ≥ N*_*p*_, and *t*(*N*_*p*_) attains its minimal value at *N*_*p*_. To find *N*_*p*_, we first solve 1 − 2*q*^2*N*^ = 0 with respect to *N*, and then choose from the two nearest integers (i.e., ⌊*N*⌋, ⌊*N*⌋ + 1) the one which minimizes *t*_*N*_. □

### Remark A.1.

Note that, if *N ≥* 1 and *pN* → 0, then for *t*(*N*) in (5) it holds

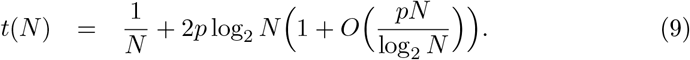

To see this, it suffices to use the following bounds

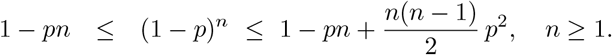

Proof of proposition 2.2. <u>Step 1 (fixed points).</u> Define

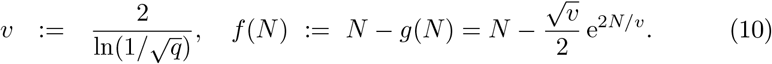

Then derivative 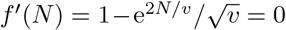 which is equivalent to 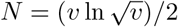. Note that *f*^*′*^(*N*) → − ∞ as *N* → ∞. Moreover, *f*^*′*^(0) *>* 0 since 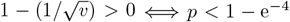 which is satisfied for any *p* ∈ *A*. Therefore, *f* attains maximal value at

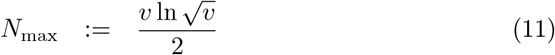

and

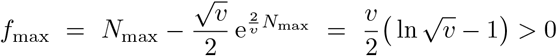

since 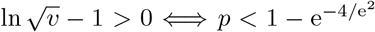. On the other hand, 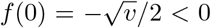 and *f* (*N*) → −∞. Consequently, *f* has two zeroes: *N*_*p*_ ∈ (0, *N*_max_) and *Ñ*_*p*_ ∈ (*N*_max_, ∞). The latter means that *g* has two fixed points.

<u>Step 2 (minimizer).</u> In this step, we show that *N*_*p*_ from Step 1 is the minimizer for *t*(*N*) given in (2). By (3),

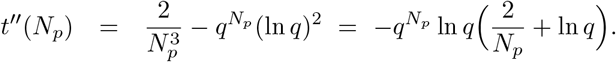

Hence,

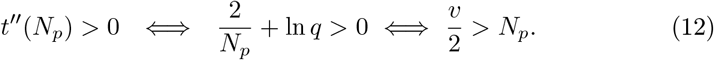

From Step 1 (see (11)) it follows that 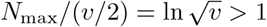, i.e. *v/*2 ∈ (*0, N*_max_). Therefore, (12) holds if and only if *f* (*v/*2) *>* 0. The latter reads as (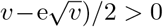 and is equivalent to 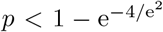 showing that *N*_*p*_ (being the critical point of *t*) is indeed the announced minimizer. Finally, note that the above analysis also implies that *Ñ*_*p*_ from Step 1 is the maximizer of *t*(*N*) which affirms the uniqueness of the minimizer. □

### Remark A.2.

One can also show that *p ↦ N*_*p*_ is strictly decreasing and continuous on *A*. However, the latter properties seem to be of less importance and we omit the details.

## B Appendix. Tables

In the tables below, the following information is provided:

- Column ‘*N*_*p*_’ shows an optimal sample size corresponding to *p* ranging in an interval given in the column ‘Range of *p*’.
- Column ‘Range of 100*G*_*p*_’ shows an average gain (as defined in the main body of the paper) per 100 individuals corresponding to values of *p* and *N*_*p*_ given in the two leading columns. The highest gain corresponds to the lowest *p* in the corresponding interval. For example, in Table 1, the first line should be interpreted as follows: for *p* ∈ [0.1865, 0.2000], optimal sample size *N*_*p*_ is equal to 2; if *p* = 0.2000, then average gain per 100 individuals 100*G*_*p*_ is equal to 16.1782; if *p* = 0.1865, then 100*G*_*p*_ = 14.000; for intermediate values of *p*, the value of 100*G*_*p*_ lies in [14.0000, 16.1782].

## Acknowledgment

We would like to thank two anonymous referees for insightful and constructive comments, which helped to improve the preliminary version of the paper.

## Footnotes

† The notations for a short reference of GT schemes come from Dorfman (**D**); Halving (**H**). **A** reflects the most naive and straightforward option.

‡ the discussed appearance of entropy in formula (6), in fact, is a simple conclusion following from Shannon’s Coding Theory; a bit more on the connections with that theory can be found in the Appendix H of the Supplementary Material of the reference [27]

§ According to Wikipedia [9], ‘In Israel, researchers at Technion and Rambam Hospital developed a method for testing samples from 64 patients simultaneously, by pooling the samples and only testing further if the combined was positive. Pool testing was then adopted in Israel, Germany, Ghana, South Korea, Nebraska, China and the Indian states Uttar Pradesh, West Bengal, Punjab, Chhattisgarh, and Maharashtra.’ Also see ‘List of countries implementing pool testing strategy against COVID-19’ therein.

